# Functional Reorganization across Four Episodes in Bipolar Disorder

**DOI:** 10.1101/2024.09.03.24312957

**Authors:** Xiaobo Liu, Bin Wan, Xi-Han Zhang, Ruifang Cui, Siyu Long, Ruiyang Ge, Lang Liu, Jinming Xiao, Zhen-Qi Liu, Jiadong Yan, Ke Xie, Meng Yao, Xin Wen, Sanwang Wang, Yujun Gao

## Abstract

Bipolar disorder (BD) is a heterogeneous psychiatric condition encompassing various episode states, including manic (BipM), depressive (BipD), mixed (mBD), and remission (rBD). Cumulative evidence has revealed that few BD episodes exhibit brain functional connectome changes; however, these findings remain fragmentary, and a systematic understanding of these functional alterations among all episodes is still lacking. Here, we firstly aimed to investigate how these episodes are differentially represented along the sensory-association axis, which constitutes the primary functional organization spectrum of the human cortex. We found expansion in sensory regions with compression in association regions in BipM, BipD and rBD as well as expansion in visual regions and prefrontal regions, compression motor regions, and precuneus regions for mBD compared to healthy controls. By examining outward and inward activity propagation in association regions, we observed less frequent flows in BipM and BipD, clarifying that association may be dominant in functional reorganization. mBD showed more frequent inward flows than other episodes in both sensory and associations, emphasizing information exchange between these two anchors. By evaluating the network integration and segregation, we observed an increase in functional integration alongside a decrease in functional segregation for unipolar episodes and the greatest functional integration for mBD. Furthermore, clinical relevance analysis suggests that emotional changes were mainly related to association functional reorganization, which may be used as specific and sensitive biomarkers to detect mood changes in different episodes. Finally, the episode representations were spatially correlated with distribution of serotonin transporter, gamma-aminobutyric acid type A receptor, Alpha-4-Beta-4 nicotinic acetylcholine receptor, layer 4 and layer 5 excitatory neurons. This study demonstrates functional reorganization as a biomarker as well as a uniform and simplified framework of neural phenotype to quantify mental abnormality of different BD episodes systematically.

## Introduction

Bipolar disorder (BD) is a psychiatric condition characterized by extreme mood fluctuations, presenting as manic (BipM), depressive (BipD), mixed episodes (mBD), and remission episode (rBD) (Goes, 2023; Grande et al., 2016a; Ljubic et al., 2023; Phillips & Kupfer, 2013). The clinical symptoms of dynamic episodes, such as emotional imbalances, may be associated with various cognitive and behavioral abnormalities and brain function (Grande et al., 2016b; Nierenberg et al., 2023). The current measurement of brain functional alterations primarily relies on functional magnetic resonance imaging (fMRI) (Liu et al., 2024b; Nan et al., 2024; Waller et al., 2021), which detects the regional blood-oxygen-level-dependent (BOLD) signal across continuous time points and offers the spatiotemporal dynamics related to brain function. Resting-state fMRI studies have indicated that emotional dysregulation and cognitive impairments in individuals with BD are linked with disrupted inter-regional connectivities (Liu et al., 2024a; Thermenos et al., 2010). Utilizing graph theory, studies have found that BD is associated with alterations in brain modularity (Jiang et al., 2025; L. Zhang et al., 2021). However, these findings remain fragmentary while functional reorganization in all BD episodes has been incompletely understood (Northoff & Hirjak, 2024; Rihmer & Kiss, 2002; Russo et al., 2020).

Brain organization emphasizes the spatial patterns of integrating and segregating inter-regional information (Margulies et al., 2016), such as functional network communities (Yeo et al., 2011) and continuous gradients of functional spectrum (Vos de Wael et al., 2020). Functional gradients provide a new insight into smooth transitions between cortical regions rather than separate communities, representing the low dimensionality of the regional functional connectome (Margulies et al., 2016). Furthermore, gradients in brain organization have been studied in multiple psychiatric conditions and demonstrate high sensitivity and robustness (Hong et al., 2020; Huntenburg et al., 2018). Functional gradients provide a robust framework for investigating brain reorganization across BD episodes, enabling the characterization of continuous spatial variability across episodes within the global sensory-association axis, rather than focusing solely on localized regional changes. Furthermore, the primary functional gradient in health have been found closely related to neurobiological substrates, including cortical morphology, superficial white matter microstructure, structure-function coupling (Vázquez-Rodríguez et al., 2019), and genetic patterns (Xia et al., 2022), highlighting the microstructural and cellular underpinnings of the sensory-association axis of cortical functional organizations (X.-H. Zhang et al., 2024). Understanding how the sensory-association gradient is distorted across BD episodes may reveal biologically tractable targets for treatment and etiological studies.

Extending the low-dimension representation approach, advances in dynamic information flow methods (e.g., regression dynamic causal model (rDCM) (Frässle et al., 2021) ) have further clarified directional interactions between sensory and association regions, elucidating dominant dynamics among functional regions. For instance, via rDCM and resting state fMRI, previous study revealed temporal lobe epilepsy showed atypical signal flow among various functional reorganization regions (Xie et al., 2024). Even though low-dimensional representation captures the continuous functional variations across the cortical surface, brain functional connectome is highly dimensional, hierarchically organized and support nested segregation and integration across multiple spatial scales (Tucker & Luu, 2023; R. Wang et al., 2021). While high-dimensional approaches reveal the hierarchical organization and topographical patterns among different brain regions (Yeo et al., 2011), further emphasizing balance between functional integration and segregation as a uniform framework to understand functional reorganization and cognition flexibility (Chang et al., 2023; R. Wang et al., 2021). A recent study has developed the Nested Spectral Partitioning (NSP) method (R. Wang et al., 2021) to determine the hierarchical functional reorganization based on segregation and integration balance from the whole high-dimension human networks (Sporns, 2013).

Together, functional reorganization, especially functional integration and separation offers a unified and simplified representation of neural phenotype to quantify mental abnormality of different BD episodes systematically. Functional integration reflects enhanced coordination between brain regions, supporting the execution of higher-order cognitive tasks, while functional separation ensures the independence of brain regions, maintaining sensory processing and basic functional stability (Capouskova et al., 2023; Cohen & D’Esposito, 2016; Numssen et al., 2021; Yeo et al., 2015). Disruptions in this balance impair the ability to switch between cognitive tasks or emotional states, exacerbating deficits in cognitive flexibility (Elvsåshagen et al., 2013; Grande et al., 2016a; McIntyre et al., 2020; Phillips & Kupfer, 2013; Rihmer & Kiss, 2002). For instance, excessive integration within the default mode network (DMN) may enhance self-referential thinking, manifesting as rumination and hyper-introspection during depressive episodes (Elvsåshagen et al., 2013; Liu et al., 2024b) while reduced integration between the central executive network and other functional networks during manic episodes can lead to impulsive behaviors and impaired executive function (Hu et al., 2023). Also, cumulative evidence has shown vulnerability of BD was associated with resting-state cohesiveness of sensorimotor network and resilience of patients was linked to integration of the default mode network (Doucet et al., 2017). Notably, these functional reorganizations in BD are underpinned by significant molecular and biological mechanisms (Poletti et al., 2017). Although these findings highlight the critical role of functional imbalances between primary sensory and higher-order cognitive networks in BD, the exact mechanisms by which these abnormalities serve as a shared etiology across different episodes remain unclear. Developing a unified model to quantify these brain functional changes is essential for advancing cross-episode diagnostics and comorbidity-targeted therapies.

Therefore, in the present study (**Figure 1**), we first examine the functional reorganization along the sensory-association axis by contrasting this primary functional gradient between BD and healthy controls (HC). We delineate distinct reorganization patterns among various episodes by further comparing these functional reorganization measures across rBD (*n* = 37), mBD (*n* = 38), BipD (*n* = 42), BipM (*n* = 38) and HC (*n* = 35) groups. Then, the directional information flows in the reorganized sensory-association axis of different BD episodes are probed via regression dynamic causal modeling (rDCM) (Frässle et al., 2021), and the separate integration and segregation in the same axis are decoded via nested spectral partitioning (NSP) (Chang et al., 2023). Finally, the reorganization patterns are compared with clinical symptoms and normative maps of cellular and receptor profiles, to investigate the symptomatic and molecular associations.

**Figure 1.**
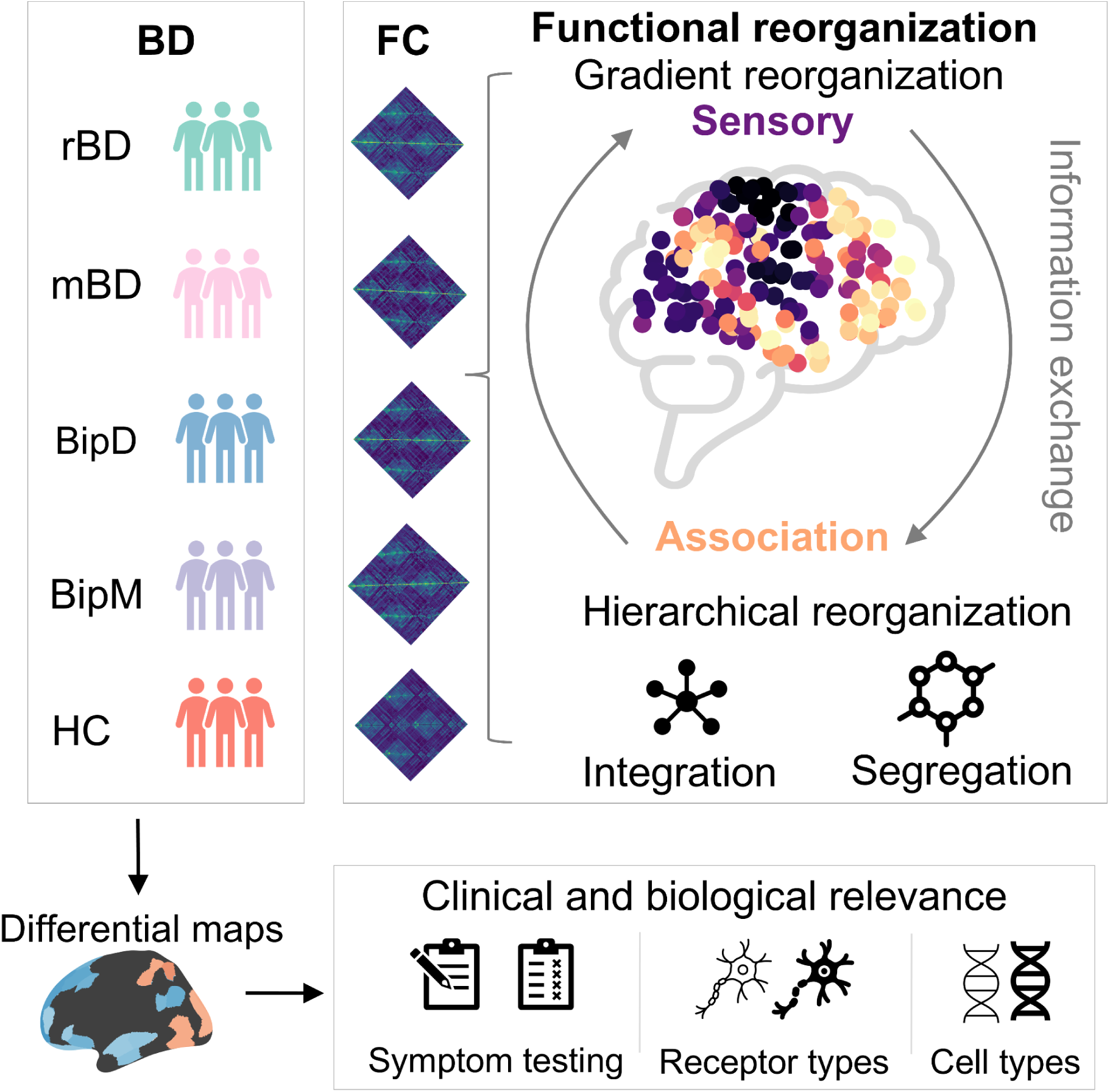
Overall framework in this study is to understand functional reorganization of four episodes of BD, including rBD, mBD, BipD, BipM and HC. Here, in this study, we firstly delineate functional reorganization via low dimension representation along the cortex and emphasize information exchange between sensory and association regions. Next, we build nested spectral partitioning (NSP) to summarize functional reorganization across spatial scale. Finally, we associated the functional reorganization patterns with clinical and biological relevance, including receptor and cell types. Note that rBD, mBD, BipD, BipM represent remission episodes,mixed episodes, depressive episodes and manic episodes respectively.

## Results

### Functional reorganization across various episodes

We applied diffusion map embedding (Coifman & Lafon, 2006; Vos de Wael et al., 2020) to cortex-wide functional connectomes derived from resting-state fMRI for each individual and investigated the alteration of functional organization gradients for each episode state compared to healthy controls. **Figure 2a** shows the variability of the functional gradient across various episodes.The average principal functional gradient maps across rBD, BipD, BipM and HC demonstrated a continuous spatial transition from sensory regions to association regions in all groups (**S-Figure 2**), accounting for 15.00 % ± 0.70%, 15.14% ± 0.92%, 15.30% ± 0.78% and 15.14% ± 0.83% of total variance respectively and aligning with previous study (Margulies et al., 2016). Notably, for 3 unipolar episodes, we found rBD, BipD, and BipM exhibited markedly higher gradient values in sensory regions and significantly lower gradient values in association regions (all *p_FDR_ <* 0.05, shown in **Figure 2b**)). The common reorganization regions across the three unipolar episodes included the visual network, dorsal attention network posterior, orbitofrontal cortex in limbic, lateral prefrontal and medial prefrontal in the control network, cingulate cortex in the control network according to Yeo atlas (Yeo et al., 2011). More specifically, those networks included temporal regions, parietal regions, prefrontal regions, precuneus/posterior cingulate cortex, dorsal prefrontal cortex/medial prefrontal cortex, and visual regions (**Figure 2c**). Note that we didn’t observe any statistically significant relationship among the three episodes after correction (**Supplementary S-Figure 5).** The specific functional reorganization regions for various unipolar episodes were displayed in **Supplementary S-Figure 3** and **S-Figure 4**. Those results showed functional reorganizations were similar across all episodes and implied it as a common principle to reveal potential shared mechanisms for unipolar episodes. Next, we further compared the functional reorganization of mixed episodes with unipolar episodes. mBD exhibited markedly higher gradients in motor regions and precuneus regions of association anchor, and lower gradient values in visual regions and prefrontal regions of association anchor, shifting from sensory toward association anchor. To further clarify the annotation of functional reorganization, we linked those cortical maps to NeuroSynth (Yarkoni et al., 2011). We chose 20 terms from (Margulies et al., 2016) and associated them with functional reorganization patterns (shown in **S-Figure 4)**. mBD reorganization patterns as well as three unipolar episodes were especially related to high-order cognition (e.g., emotion inhibition, autobiographical memory, and social cognition), confirming three unipolar episodes and mBD showed similar brain functions. In all, we observed that expansion in sensory regions and compression in association were general functional reorganization principles of three unipolar episodes. Expansion in visual regions and prefrontal regions, compression motor regions, and precuneus regions were functional reorganization for mixed episodes, in contrast to HC. However, the three unipolar episodes did not differ from each other.

**Figure 2.**
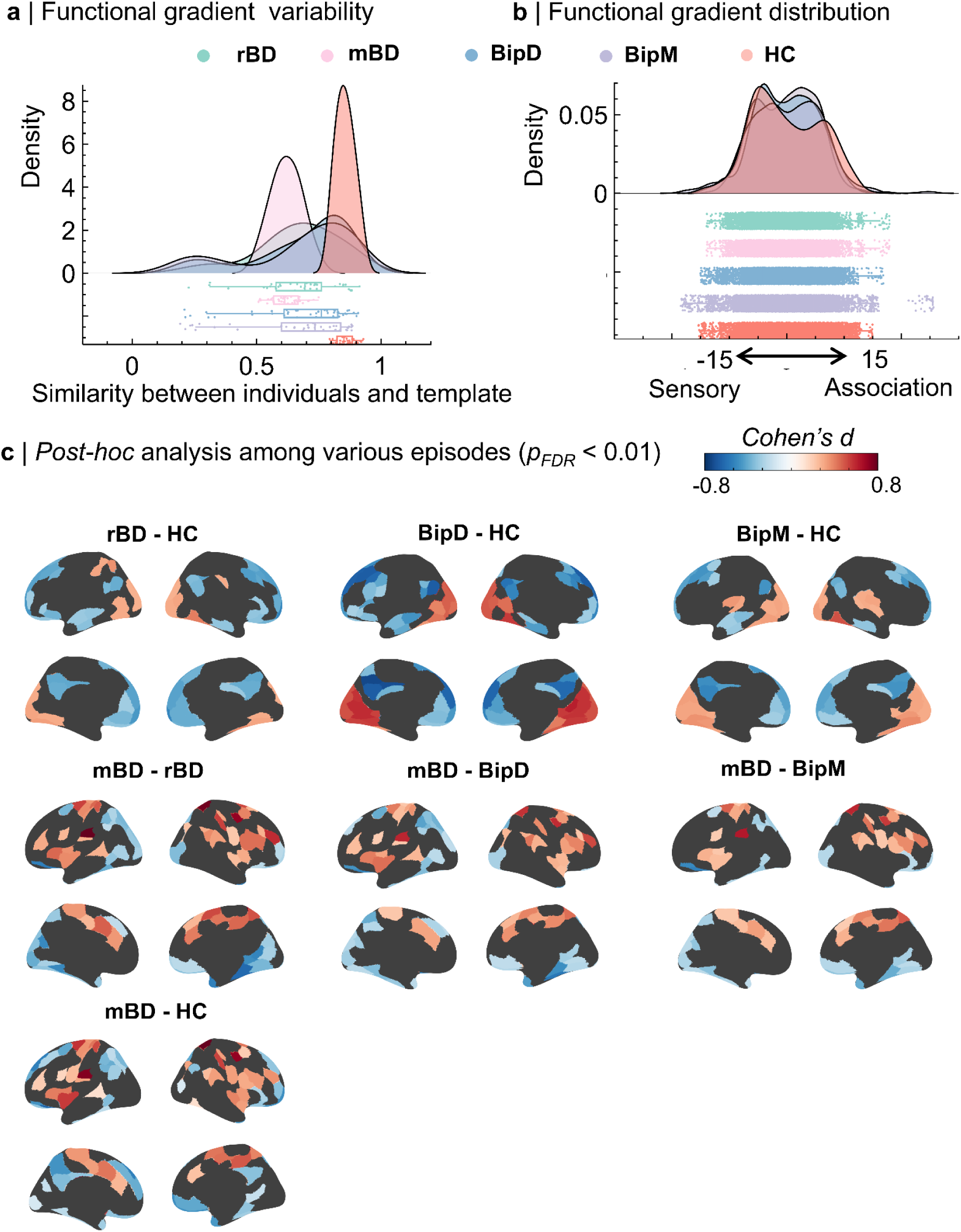
Functional reorganization in flow dimension space in various episodes, including rBD (N = 37), mBD (N = 38), BipD (N = 42), BipM (N = 38) and HC (N = 35) groups. **a.** Variability of functional gradient across different BD episodes. Note that variability represents the similarity between individual functional gradient maps and templates. **b.** Distribution of functional gradients from various episodes. **c.** The comparison of functional gradients among rBD, mBD, BipD, BipM and HC.

**Figure 3.**
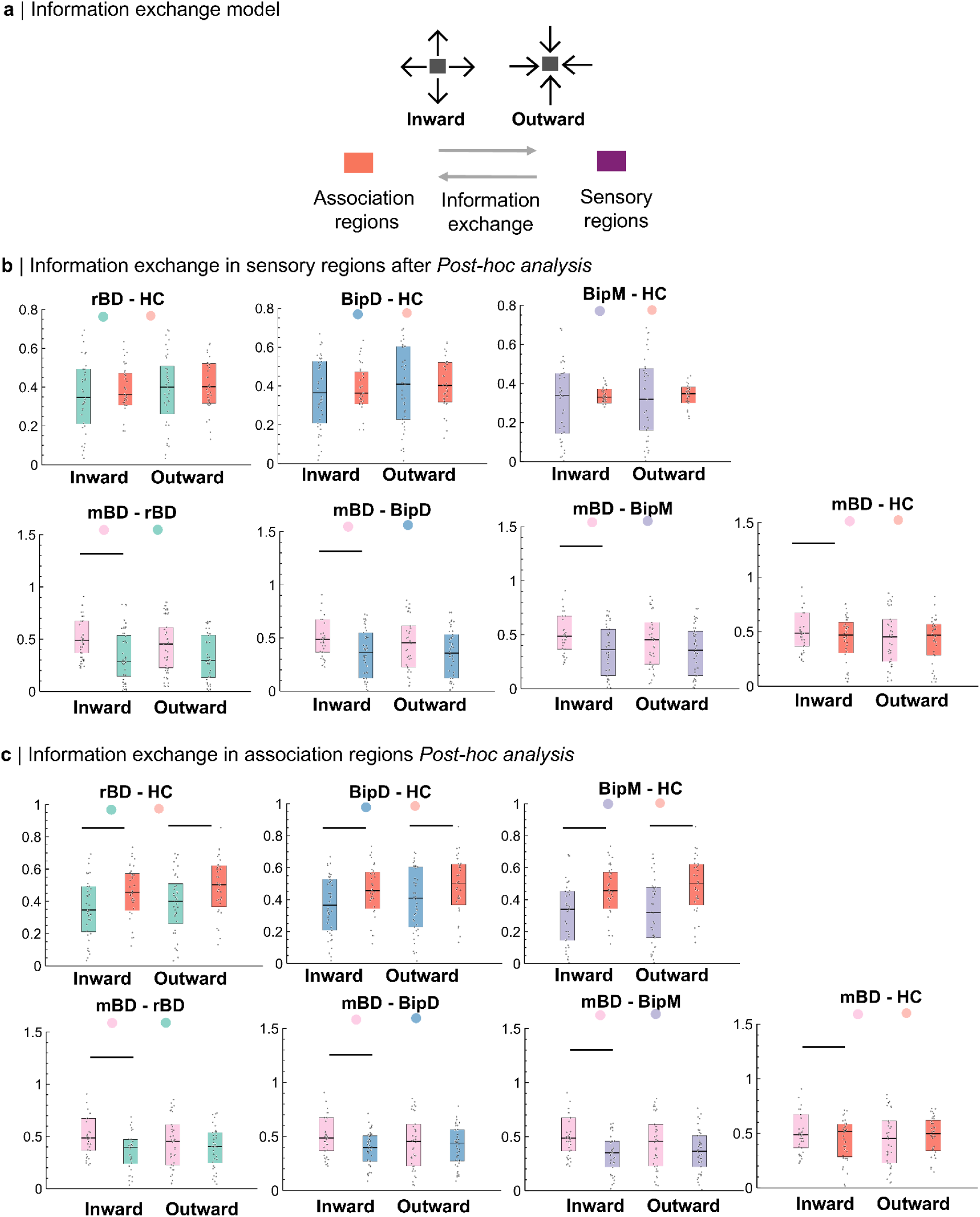
Outward and inward degree in the sensory and association regions**. a.** Outward and inward of functional reorganization regions across rBD, mBD, BipM and BipD via one-way ANOVA and *post-hoc* analysis. Note that the arrows represent information exchange among sensory regions and association regions via inward and outward degree. **b.** The abnormal information flows in various episodes in sensory regions (*P_HSD_ < 0.05*). Note that 一 means significant after *post hoc* analysis and HSD. **c.** The abnormal information flows in various episodes in association regions (*P_HSD_ < 0.05*).

**Figure 4.**
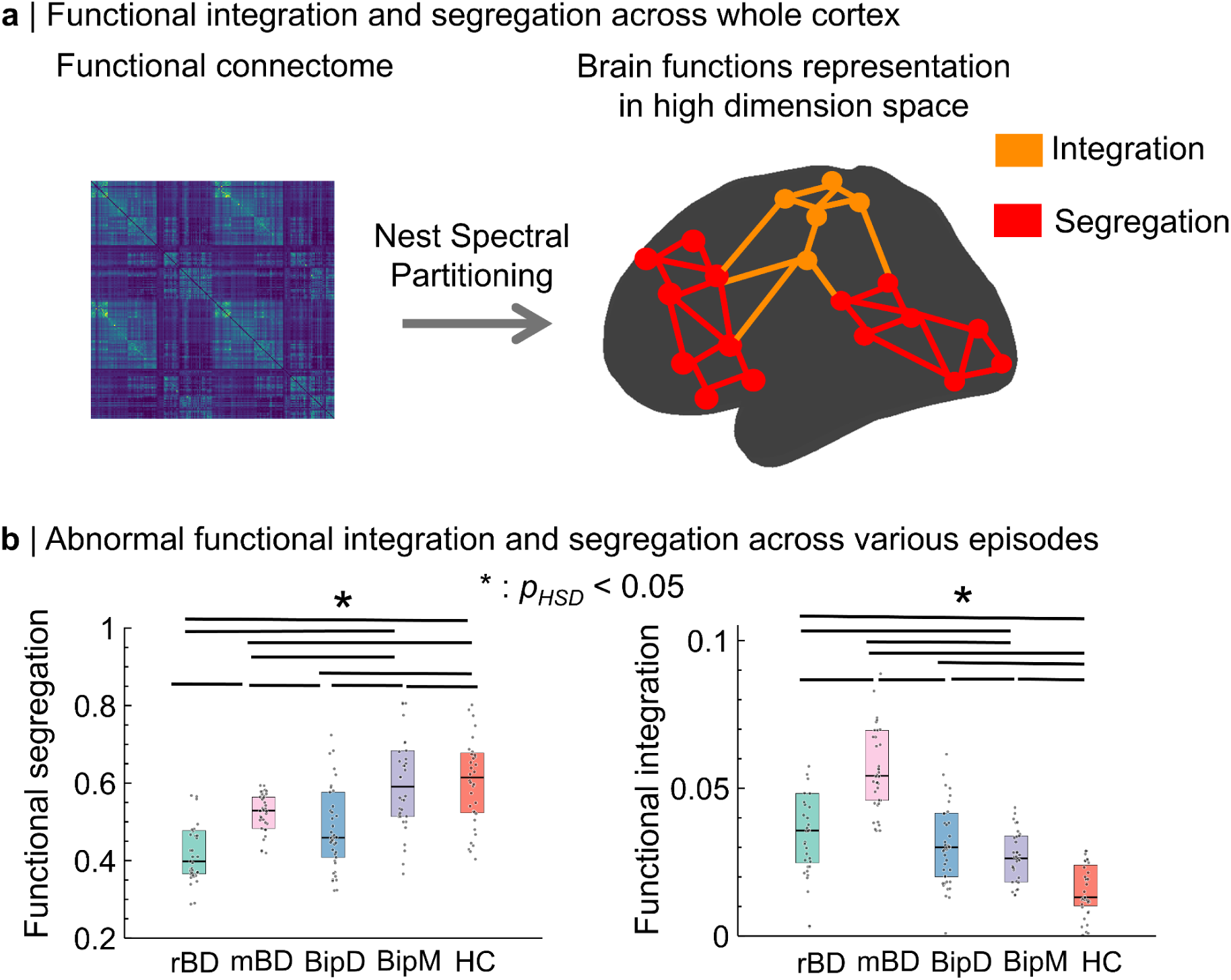
Functional separation and integration across the whole cortex among rBD, mBD, BipD and BipM. **a.** Whole cortex functional integration and functional segregation. **b.** Whole cortex functional integration and functional segregation among rBD, mBD, BipD and BipM (* means *p_HSD_ < 0.05* after *post-host* analysis).

**Figure 5.**
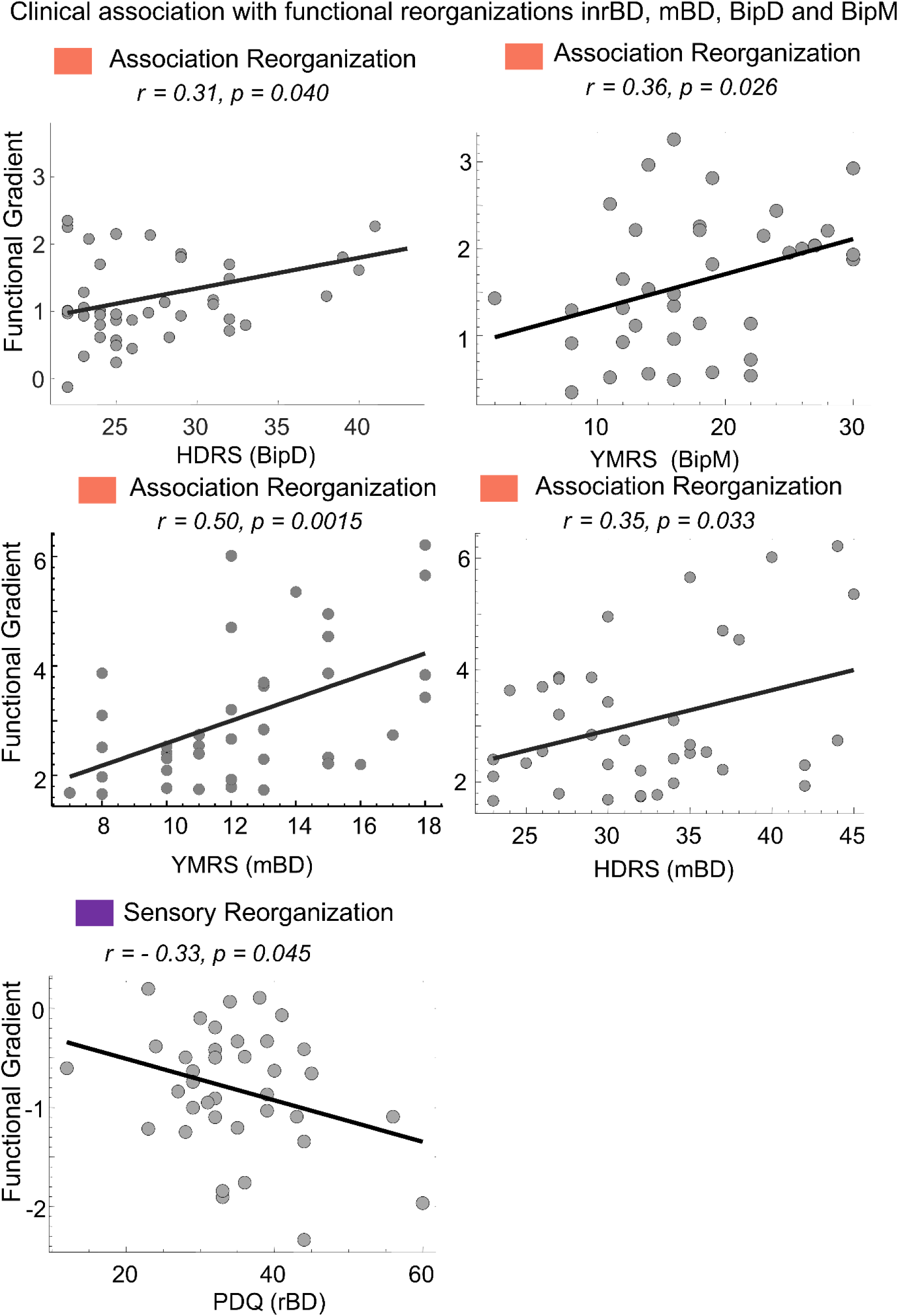
Functional reorganization in association as well as sensory regions and clinical symptoms across various BD episodes. Association reorganization was positively related with HDRS both in BipD (*r =0.32, p = 0.038*) and mBD (*r =0.50, p = 0.0015*) and with YMRS both in mBD ( *r = 0.35, p = 0.033*) and BipM ( *r =0.43, p = 0.006*). Also, the sensory reorganization in rBD was negatively related with PDQ scores ( *r = - 0.33, p = 0.045*). Note that HDRS, Hamilton Depression Rating Scale; YMRS, Young Mania Rating Scale; PDQ, Perceived Disability Questionnaire .

### Information flow in reorganization regions across various episodes

After identifying functional reorganization regions in various episodes, we further explored how information flows clarified dominance between sensory and association regions (**Figure 3a**) and implemented rDCM to estimate the cortex-wide effective connectome for each participant. Here, we calculated outward and inward degree centrality in both sensory and association regions to represent information flow across regions in both unipolar episode and mixed episodes. To simplify the analysis, we averaged outward and inward degrees in the significant sensory and association regions respectively and focused on the significant comparison in **Figure 2b**. Via one-way analysis of variance (ANOVA), we found there was a significant difference in outward (*F_(4,186)_ =* 5.58, *P =* 0.0003) and inward (*F_(4,186)_ =* 4.34, *P =* 0.0022) degrees in both association regions sensory regions between rBD/mBD/BipD/BipM and HC (**Figure 3b** and **Figure3c**). Specifically, the *post hoc* analyses revealed that the both average inward degree and outward degree of rBD, mBD, BipD and BipM is significantly higher in association regions, compared to HC ( *P_HSD_ <* 0.05). However we didn’t find any statistical difference in both average outward and inward degrees in sensory regions between episodes and healthy controls ((*p_HSD_ >* 0.05), indicating the information flow differences were specific to association regions. Then, we found that the average intward degree in both association regions and sensory regions for rBD, BipD, and BipM, compared to mBD (*P_HSD_ <* 0.05). Those results further clarified association regions dominated the functional reorganization along cortex continuously in unipolar episodes. while both association regions and sensory regions for mixed episodes.

### Functional integration and separation in high dimensional approaches among various episodes

Although functional gradients offer a perspective to detect both integration and segregation along the cortex continuously, high-dimensional approaches summarize the hierarchical organization and topographical patterns across various spatial scales. Therefore, we explored the separate functional integration and segregation across various episodes via NSP (**Figure 4a**). We observed that the first component of NSP was significantly positive related to the functional gradient map (*r = 0.51*, *p <* 0.001) in **Supplementary S-Figure 8**, suggesting NSP revealed similar information as functional gradient. To further explore association between NSP values and functional reorganization, integration scores defined by NSP were positively correlated with sensory reorganization (*r* = - 0.19, *p* < 0.05) and segregation scores were negatively correlated with association reorganization regions (*r* = 0.20, *p* < 0.05) in **Supplementary S-Figure 9**. This suggests that the reorganization of sensory regions is more related to functional integration, while the reorganization of association regions is more associated with functional segregation. Functional separation significantly increased (*F_(4,186)_* = 15.24, *p <* 0.001 ) while functional integration significantly decreased from rBD, BipD and BipM (*F_(4,186)_* = 34.76, *p* < 0.001) (**Figure 4b**). For unipolar episodes, *Post-hoc* analyses showed rBD had significantly lower functional separation than BipD, BipM and HC while BipD had significantly lower functional separation than BipM and HC (*p_HSD_ <* 0.05). In contrast, rBD had significantly higher functional integration than BipD, BipM and HC, BipD had significantly higher functional integration than BipM and HCand BipM also had higher functional integration than HC (*p_HSD_ <* 0.05). These results further emphasize that the dynamic changes in integration and segregation during functional reorganization in unipolar episodes may reflect a progression of functional imbalance from rBD to BipD, BipM, and finally to HC. For mixed episode, we found functional segregation in mBD was significantly higher than rBD and BipD but significantly lower than BipM and HC, while its functional integration was significantly higher than other episodes (*p_HSD_ <* 0.05). In summary, these findings highlight integration and segregation as holistic principles for understanding functional reorganization in BD. They suggest that from rBD to BipD and BipM, the capacity for functional integration progressively weakens, indicating a gradual decline in integrative capacity along this trajectory in unipolar episodes. Also, mixed episode showed the strongest functional integration capacity across all episodes.

### Functional reorganization and clinical symptom in various episodes

Next, we probed the clinical association with neuroimaging findings (**Figure 5)** to further confirm functional gradient as biomarker to quantify functional abnormality of various episodes of BD. Specifically, the analysis also revealed positive correlation between the HDRS score and the reorganization of the association areas in BipD (*r* = 0.32, *p* = 0.038), and between the YMRS score and reorganization of the association areas (*r* = 0.43, *p* = 0.006) in BipM. For mBD, sensory reorganization (visual regions) was negatively associated with YMRS (*r* = - 0.36, *p* = 0.003) and association reorganization (precuneus regions) was positively associated with HDRS (*r* = 0.35, *p* = 0.033). However, we didn’t find any significance between the functional reorganization in association areas and YMRS in BipD( *r = - 0.18*, *p = 0.26*), HDRS in BipM( *r = 0.17*, *p = 0.34*) and both YMRS ( *r = 0.1*, *p = 0.87*) and HDRS ( *r = - 0.21*, *p = 0.22*) in rBD (**Supplementary S-Figure 10)**. There is not any significance between sensory reorganization and YMRS and HDRS in various episodes ( *p > 0.05*). These results suggest that extreme mood changes are mainly related to functional reorganization in association regions, which may be used as specific and sensitive biomarkers to detect mood changes in different episodes . Also, we observed a significantly negative correlation between the PDQ score and the functional reorganization of sensory areas in rBD (*r = - 0.33, p =* 0.045). More specifically, extension of functional gradient in sensory areas may exacerbate difficulties or impairments in perception for these patients. These findings confirmed clinical relevance of the functional reorganization for various BD episodes and further revealed that functional reorganizations may be caused by emotion alternation.

### Molecular underpinnings functional reorganization

Next, we further tested biological association of functional reorganization patterns and focused on common regions across 4 episodes. We conducted spatial correlation analyses between the molecular or cellular maps and functional reorganization maps for each episode, including unipolar episodes and mixed episodes. The molecular and cellular maps are from Neuromaps (Markello et al., 2022; X.-H. Zhang et al., 2023), which provides normative maps for various neurotransmitter systems and cellular systems. Here the spatial correlation reflects which receptor and cellular types association aligned spatially with the sensory-association reorganization axis across various episodes. Specifically, negative association values showed higher cell or neurotransmitter density more related to sensory reorganization while positive correlation showed more related to association reorganization. Notably, **Figure 6a** showed that reorganization of sensory areas in unipolar episodes was correlated with Paired box gene-cell (Pax6), Synuclein (Sncg), Astrocyte (Astro) and Layer 5 thick-tufted (L5IT) and reorganization of association areas was correlated with L4IT (Layer 4 thick-tufted). The functional reorganization of association for mixed episode was associated with Layer 5 Extratelencephalic neurons (L5-ET), Endothelial cells (Endo), Layer 6 Intratelencephalic neurons expressing Car3 (L6-IT-Car3) and Layer 2/3 Intratelencephalic neurons (L2/3 IT). These associations suggest that specific genes and cell types may have distinct contributions and mechanisms in the process of functional reorganization and adaptation in different brain regions. This indicates that functional changes in different brain areas may depend on specific molecular and cellular characteristics. At the cellular level, we also found that reorganization of association areas in unipolar episodes was correlated with the Serotonin Transporter (5 HTT), Dopamine Transporter (DAT), Gamma-Aminobutyric Acid Type A Receptor (GABAa), whereas reorganization of sensory areas was correlated with the Alpha4Beta4 Nicotinic Acetylcholine Receptor Subtype (α4β4), Cannabinoid Receptor Type 1 (CB1) and Vesicular Acetylcholine Transporter (VAChT) (shown in **Figure 6b**). The association functional reorganization was associated with 5-Hydroxytryptamine receptor 2A (5HT2a) and 5-Hydroxytryptamine receptor 4 (5HT4) and sensory functional reorganization was associated with Dopamine receptor D1 (D1) and Norepinephrine transporter (NET) in mixed episodes. These findings suggest that the functional reorganization of different brain regions may rely on distinct neurotransmitter and receptor signaling mechanisms, which in turn influence their functional performance under specific pathological or adaptive conditions. Together, these findings suggest that the functional reorganization pattern across various BD episodes aligned topographically with the cortical distribution of chemical neuromodulator systems and implied their potential molecular biomarker.

**Figure 6.**
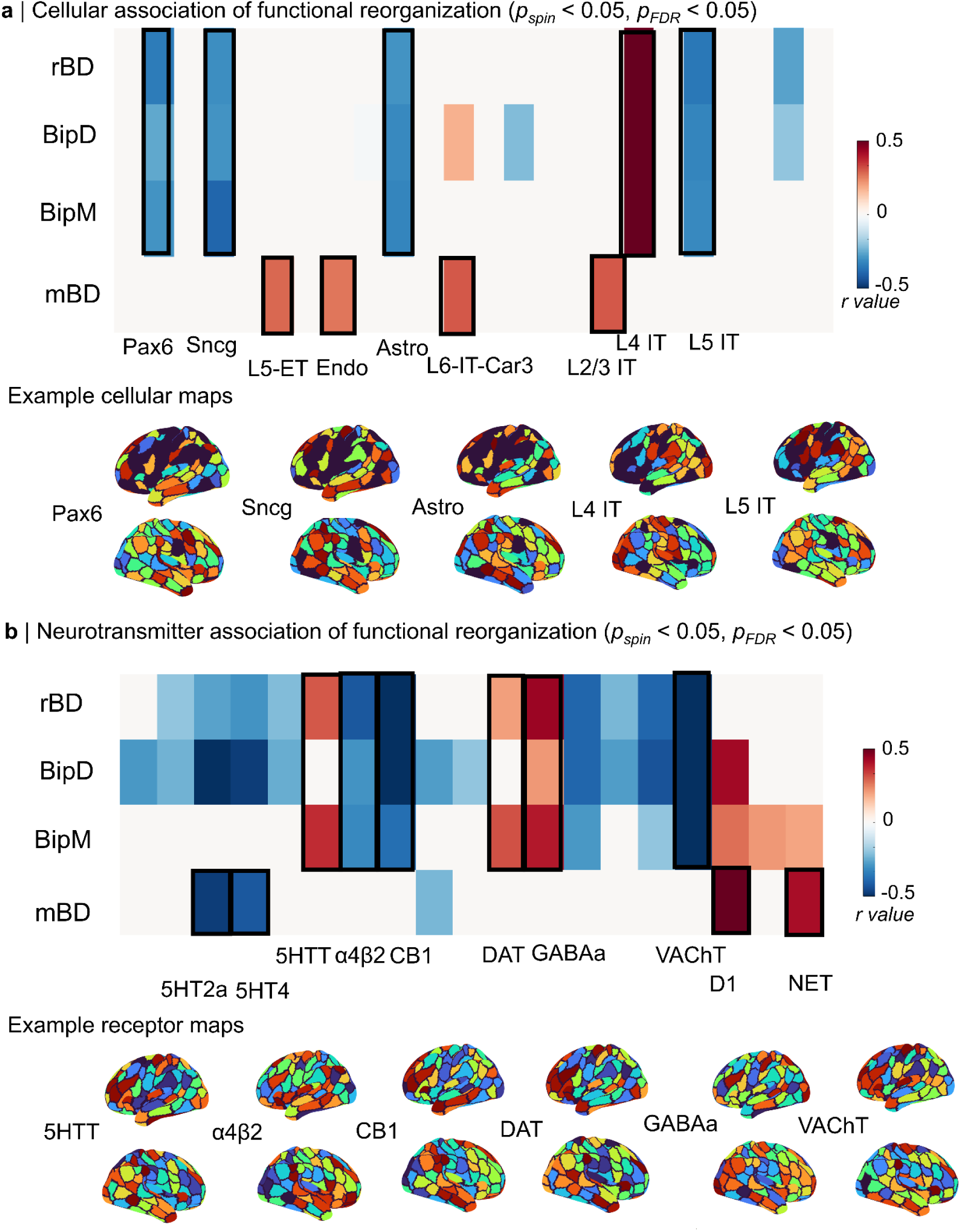
Molecular mechanism of functional reorganization among rBD, mBD, BipD and BipM (*p_spin_* < 0.05, *p_FDR_* < 0.05). **a.** Cellular mechanism of functional reorganization. We found sensory reorganization correlated with Pax6, Sncg, Astro, L4IT and L5IT and association reorganization correlated with L4IT in unipolar episodes. And association functional reorganization in mixed episodes was associated with L5-ET, Endo, L6-IT-Car3 and L2/3 IT. **b.** Receptor mechanism of functional reorganization. Specifically, association reorganization correlated with 5HTT, DAT, GABAa and sensory reorganization correlated with α4β4, CB1, and VAChT for unipolar episodes. The association functional reorganization was associated with 5HT2a and 5HT4 and sensory functional reorganization was associated with D1 and NET in mixed episodes. Note that Pax6, Sncg, Astro, L4IT, L5IT, L5-ET, Endo, L6-IT-Car3, L2/3 IT, 5HTT, DAT,GABAa, α4β4, CB1, VAChT D1 and NET means Paired box gene, Synuclein, Astrocyte, Layer 4 thick-tufted, Layer 5 thick-tufted,Layer 5 Extratelencephalic neurons, Endothelial cells, Layer 6 Intratelencephalic neurons expressing Car3, Layer 2/3 Intratelencephalic neurons, Serotonin Transporter, Dopamine Transporter, Gamma-Aminobutyric Acid Type A Receptor, Alpha4Beta4 Nicotinic Acetylcholine Receptor Subtype, Cannabinoid Receptor Type 1 and Vesicular Acetylcholine Transporter, dopamine receptor and Norepinephrine Transporter.

## Discussion

In the present study, we uncovered functional connectome reorganization characteristics of multiple episodes in BD. Specifically, we identified a dichotomy in reorganization: sensory regions expanded and association regions compressed in unipolar episodes. Whereas, functional reorganization in mBD showed predominantly from sensory to association regions. Regional activity propagation revealed reduced outward and inward flows in association regions during unipolar episodes, highlighting their dominant role in functional reorganization. In contrast, mBD exhibited more frequent inward flows, reflecting enhanced sensory-association interactions. A network integration-segregation analysis showed increased integration and reduced segregation in unipolar episodes, with mBD displaying the highest integration levels across all episodes. Furthermore, clinical relevance analysis suggests that emotional changes were mainly related to association functional reorganization, which may be a specific and sensitive biomarker for different episodes. Molecular correlates of these reorganizations included serotonin transporter, gamma-aminobutyric acid type A (GABA_A) receptor, alpha-4-beta-4 nicotinic acetylcholine receptor, and cellular profiles of Layer 4 and Layer 5 thick-tufted cells.

The expansion of sensory regions and compression of association regions suggests a significant reconfiguration of brain network architecture in unipolar episodes. Previous studies have also found compression of functional gradients in the association regions of BD (Lin et al., 2023) and gradients in sensory regions (Lei et al., 2023). Functional reorganization from sensory to association in mBD further revealed there may be greater reliance on sensory regions for processing external information and self-regulation, while association regions may be due to heightened emotional conflict and cognitive burden, reducing their functional influence as emotional fluctuation (Berger et al., 2007; Citrome et al., 2024; Viviani, 2013; Yatham et al., 2021). It suggests that association may be a common marker for network reorganization in psychiatry (van den Heuvel & Sporns, 2019) such as autism spectrum disorder (Hong et al., 2019), major depression disorder (Pasquini et al., 2023), and schizophrenia (Dong et al., 2020). Our findings also suggest that such reorganization is a common marker for episodes in BD, though there is no statistical difference between unipolar episodes. The functional gradient reflects both global integration and segregation (Margulies et al., 2016). In this study, functional reorganization in three unipolar episodes indicated that sensory and motor areas, which are primarily responsible for processing external stimuli, become more prominent, while association regions involved in higher-order cognitive functions lose their functional influence (Buckner et al., 2009). Such reorganization could be a compensatory mechanism where the brain reallocates resources to maintain basic sensory and motor functions at the expense of higher-order cognitive processes. This finding is consistent with previous studies that have reported reduced connectivity within the DMN (R. Wang et al., 2022) and affective networks during acute episodes but not in remitted states of BD.

Our study also highlights the significant role of aberrant association information interaction in unipolar episodes of BD, compared with sensory regions. We found that the signal outward flow and inward flow of association regions were significantly lower in three unipolar episodes of BD compared to healthy individuals. A widely accepted mechanism underlying bipolar disorder is the early developmental disruption in brain networks that regulate emotional behavior (e.g., white matter connectivity and prefrontal pruning), leading to reduced connectivity between the ventral prefrontal network and limbic brain regions (Strakowski et al., 2012). Those regions consist of association regions and their reduced connectivity suggests a core dysfunction in BD that transcends symptomatic episodes, contributing to the cognitive and emotional dysregulation observed in BD (Wu et al., 2023). Also, we found inward flow in both sensory regions and association regions in mBD was highest across all episodes. Indeed, the mBD is often characterized by extreme emotional and behavioral states, with heightened emotional conflict and cognitive burden, leading to enhanced functional integration and information exchange within association regions (Corponi et al., 2020; Mignogna & Goes, 2024). Our findings are aligned with the previous assumption and further clarified association regions were dominated in the functional reorganization across various episodes of BD.

Using large-scale functional connectivity modeling, we observed a significant increase in the brain’s integration capacity and a decrease in its segregation capacity during BipM, BipD, and rBD. While mBD had the highest integration capacity among all episodes, compared to HC. These alterations suggest a shift towards a more globally connected but less modular brain network organization in BD. A normally functioning brain at rest maintains a balance between segregation and integration, allowing for efficient communication both locally and globally. This balance is associated with the integration of primary sensory information and emotion regulation (Qin et al., 2012). Previous studies have indicated that imbalances in this functional segregation and integration may be related to functional abnormalities in various brain disorders (Chang et al., 2023). In this study, we further emphasize that the hierarchical structure of integration-segregation may underlie the emotional abnormalities and consequent behavioral anomalies observed across different BD episodes. Increased integration capacity may reflect compensatory mechanisms attempting to maintain cognitive function, while decreased segregation capacity could indicate a loss of specialized processing within distinct functional domains. This shift in network organization is consistent with findings from studies comparing functional connectivity patterns between unipolar and bipolar depression, which have shown altered connectivity in prefrontal and limbic networks (Alamian et al., 2017).

Finally, our study identifies sensory-association functional reorganization anchors across BD episodes, which are strongly associated with specific receptors and cell types. Sensory reorganization correlated with Pax6, Sncg, Astro, and L5IT, while association reorganization was linked to L4IT. At the cellular level, association reorganization was associated with 5HTT, DAT, and GABAa, whereas sensory reorganization correlated with α4β4, CB1, and VAChT. These findings highlight that the functional properties of the human brain are influenced by the diverse cell types in the cortex, with cellular spectra aligning with the macroscopic organization of functional gradients and networks (X.-H. Zhang et al., 2023). 5HT4 receptors modulate serotonin signaling, crucial for mood regulation and cognition (Bockaert et al., 2008), while 5HTT receptors regulate serotonin availability (Homberg & van den Hove, 2012). CB1 receptors, part of the endocannabinoid system, influence mood and synaptic plasticity (Viveros et al., 2007). GABA receptors modulate neuronal excitability and balance excitation and inhibition (Sears & Hewett, 2021) and mGluR5 receptors contribute to synaptic plasticity and excitatory neurotransmission (Ben-Ari et al., 2012). Our results suggest that these receptors modulate synaptic plasticity and firing patterns, impacting large-scale brain network integration and segregation. These abnormalities likely reflect the molecular basis of emotion-driven brain function and behavioral disruptions in BD. Similarly, Pax6 (Kikkawa et al., 2019), Snc (Mederos & Perea, 2019) and L5IT single-cell (Romand et al., 2011) spectra are indicative of specific cell types and their gene expression profiles, which are intricately linked to the observed changes in brain network organization. The modulation of these receptors and cells may influence the underlying mechanisms of bipolar disorder, shedding light on its pathophysiology.

## Limitation

Several limitations warrant consideration in this study. Firstly, our sample was sourced from a single site, and the relatively small sample size may constrain the reliability and generalizability of our findings. Future research will focus on multicenter studies with larger samples to address these limitations. Such an approach will enhance the external validity of our research, facilitating the extrapolation of findings to a broader patient population. Secondly, our study included both initial BipM patients and those with rBD beyond the medication washout period to gain a comprehensive understanding of the imaging features of BD. Even though we included the first episode participants in this study, it is important to acknowledge the potential impact of medications on brain function as a limitation though this is a common challenge in BD research (Wei et al., 2023), and an ideal solution has yet to be identified. Lastly, the receptor maps and cellular maps utilized in our study were derived from group-averaged results of previous research, without accounting for individual differences. Future studies will need to validate these findings by considering individual variability.

## Conclusion

In summary, our study systematically elucidates brain functional reorganizations during various BD episodes. We argued functional reorganization may serve not only as a specific biomarker for detecting mood changes across different episodes of bipolar disorder but also as a unified and simplified neural phenotype framework for systematically quantifying mental abnormalities across these episodes. The identified patterns of sensory expansion and association compression suggest a reconfiguration of brain network architecture that aligns with compensatory mechanisms and core dysfunctions in BD. The associations with specific receptors and cell spectra further highlight the molecular basis of these changes, offering preliminary evidence linking function to biology. These findings underscore the importance of integrating genetic, molecular, and functional imaging data to understand multi-level changes related to various episodes of BD.

## Method

This study was conducted following ethical approval from the Ethics Committee of Renmin Hospital of Wuhan University (Ethics Approval Number: WDRY22022-K195). Prior to study initiation, comprehensive explanations of the objectives, procedures, and precautions were provided to participants and their families, particularly for those patients who lacked decision-making capacity. Informed consent was obtained only after confirming that all participants and their families were fully informed and had a clear understanding of the study details. Subsequently, written informed consent forms were signed. The study was registered with the National Medical Research Registration Information System and the Chinese Clinical Trial Registry (Registration Number: ChiCTR2200064938).

### Clinical symptoms and cognitive assessment

The Young Mania Rating Scale (YMRS), is a clinical assessment tool designed to evaluate the severity of manic symptoms. It comprises 11 items, with scores ranging from 0 to 4 for items 1, 2, 3, 4, 7, 10, and 11, and from 0 to 8 for items 5, 6, 8, and 9 (Young et al., 1978). Clinicians assess each item based on patient responses, and the total score guides the understanding and management of manic symptoms. The Hamilton Anxiety Scale (HAMA) objectively assessed anxiety symptoms. It includes 14 items, each scored from 0 to 4, representing levels of symptom severity: None (0), Mild (1), Moderate (2), Severe (3), and Very Severe (4). Anxiety factors are categorized into somatic and psychic, and scores are analyzed accordingly (Jiang et al., 2025). The Hamilton Depression Rating Scale (HDRS) is widely used to assess the severity of depression (Hamilton, 1960). The HDRS-17 is a newer version that categorizes disease severity with lower scores indicating milder conditions and higher scores indicating more severe cases. HDRS is divided into seven factors: Anxiety/Somatization, Weight, Cognitive Disturbances, Diurnal Variation, Retardation, Sleep Disturbances, and Desperation. The Perceived Disability Questionnaire (PDQ) assesses the degree of impairment in daily life, work, social, and leisure activities. It consists of 20 items, each scored on a 5-point scale from 0 (no difficulty) to 4 (extremely difficult). The questionnaire covers various functional activities such as concentration, work completion, social interaction, and household chores.

### Imaging Dataset

We Collected data from patients diagnosed with Bipolar Disorder who sought treatment at the Department of Psychiatry and Psychology, Wuhan University People’s Hospital, from September 2021 to December 2023. Initial screening of research participants was conducted using the Chinese version of the Mini-International Neuropsychiatric Interview (MINI) (Amorim, 2000). All participants were diagnosed by two experienced psychiatrists following the Diagnostic and Statistical Manual of Mental Disorders Fifth Edition (DSM-5) criteria for BD (Association & others, 2000). Inclusion and exclusion criteria for the BipM group (Grande et al., 2016c): Age 18-45; DSM-5 diagnosis of BD, YMRS > 8, and HDRS < 7; First episode and untreated or first-time undergoing treatment. The criteria for BipD group (Tohen et al., 2012) was HDRS -17 > 12 and YMRS < 7; Inclusion criteria for rBD group: Age 18-45; DSM-5 diagnosis of BD, HDRS < 7, and YMRS < 7; Inclusion criteria for mBD group: Age 18-45; DSM-5 diagnosis of BD, HDRS > 12, and YMRS > 8; Patient-initiated discontinuation for more than 14 days. Exclusion criteria: MRI contraindications; Organic brain diseases; Other mental illnesses; History of medication or physical therapy for BipM patients; Left-handedness; Unstable physical illnesses; Substance abuse history; Pregnancy or lactation; Concurrent other brain function disorders. HCs were recruited through community, university, and Hubei Provincial People’s Hospital posters, with age and gender to the patient group; right-handedness. Exclusion criteria for HCs: MRI contraindications; Organic brain diseases; Substance abuse history; Pregnancy or lactation; Family history of neurological or psychiatric disorders. All participants had withdrawal and termination criteria: Withdrawal criteria included voluntary withdrawal of informed consent and researcher judgment of unsuitability to continue. Termination criteria involved non-cooperation leading to data invalidation, exclusion from final data analysis, and discontinuation of further investigation. MRI images were acquired using an Achieva 3T MRI scanner (GE, SIGNA Architect) equipped with a 48-channel head coil. Participants were instructed to stay awake, remain motionless, relax, and keep their eyes closed during the scanning procedure. To minimize head movement and reduce scan noise, foam padding and soft earplugs were provided. All scans were performed by two licensed MRI technicians with intermediate professional titles in the MRI room. T1_3D data sets were acquired with a maximum TR, minimum TE, a NEX of 1, a layer thickness of 2mm, and a field of view of 256 × 256 mm^2^. Scan time = 7 minutes. For rs-fMRI, a TR of 2000ms, a TE of 30ms, a FOV of 220mm × 220mm, a flip angle of 90°, a matrix of 64 × 64, a resolution of 3 × 3 × 3, a slice thickness of 36, and 240 time points were acquired. Scan time = 9 minutes.

The demographics were shown in **Table 1**. And the quality control could be seen in **S-Figure 1**.

**Table1.** Demography (HDRS means Hamilton Depression Rating Scale, YMRS means Young Mania Rating Scale,PDQ means Perceived Disability Questionnaire, HAMA means Hamilton Anxiety Scale)

|  | *Age* | *Sex(M/F)* | *handness*<br>*(R/L)* | *HDRS* | *YMRS* | *PDQ* | *HAMA* |
| --- | --- | --- | --- | --- | --- | --- | --- |
| *rBD* | 24.3±6.5 | 17/20 | 37/0 | 3.54±3.09 | 3.5±2.7 | 34.78±8.97 | 12.18±4.75 |
| *BipD* | 21.9±5.2 | 15/27 | 42/0 | 22.6±3.3 | 3.24±3.09 | 45.43±13.25 | 24.88±7.81 |
| *BipM* | 24.9±6.4 | 21/17 | 38/0 | 5.6±0.8 | 16.90±6.70 | 31.25±12.17 | 17.87±6.97 |
| *mBD* | 17.5±4.1 | 15/23 | 38/0 | 32.32±6.20 | 11.42±3.15 | ----- | ----- |
| *HC* | 26.7±7.3 | 14/21 | 35/0 | ----- | ----- | ----- | ----- |
| *F*<br>*values* | 0.0307 | 0.0054 | ----- | 321.05 | 111.61 | 15.67 | 35.67 |
| *P*<br>*values* | 0.96 | 0.98 | ----- | < 0.001 | < 0.001 | < 0.001 | < 0.001 |

### Data preprocessing and Functional connectome

For all datasets, raw DICOM files were converted to Brain Imaging Data Structure (BIDS) format using HeuDiConv v0.13.1 . The structural and functional preprocessing of both Bipolar were performed with fMRIPrep 23.0.2 (Esteban et al., 2019), which is based on Nipype 1.8.6 (Gorgolewski et al., 2011). The main anatomical data preprocessing steps include intensity normalization, brain extraction, tissue segmentation, surface reconstruction, and spatial normalization. The main functional data preprocessing steps include head motion correction, slice-time correction, and co-registration. For original preprocessing details generated by fMRIPrep. The derived functional time series were parcellated into the Schaefer 200×7 atlas and underwent a confound removal process implemented in Nilearn . The confound removal process adopted the “simple”’ strategy from (H.-T. Wang et al., 2024), including high-pass filtering, motion and tissue signals removal, detrending, and z-scoring. Functional connectivity matrices were then estimated for each subject using zero-lag Pearson correlation coefficient.

### Functional organization gradients

We initially generated cortex-wide functional connectome gradients utilizing BrainSpace (version 0.1.10; https://github.com/MICA-MNI/BrainSpace) with default parameters. Consistent with prior study (Vos de Wael et al., 2020), we retained the top 10% of weighted connections per region following z-transformation of the data to return more robust gradient loadings (Margulies et al., 2016). To capture the similarity in connectivity profiles between regions, we constructed an affinity matrix using a cosine similarity (Vos de Wael et al., 2020). We employed diffusion map embedding, a robust non-linear manifold learning technique to identify low-dimensional representations from high dimensional functional connectome for each individual (Vos de Wael et al., 2020). We extracted the first 10 components but used the first one in this study to focus on the sensory-association axis. Then individual-level gradient maps were aligned to template gradients generated from 100 unrelated healthy adults from the human connectome project database using Procrustes rotation (Wan et al., 2022) .

To estimate functional reorganization across various BD episodes, we implemented One-way ANOVA to compare the functional organization pattern between rBD/BipD/BipM and healthy control (HC) respectively with age and sex as covariates. Then False Discovery Rate (*FDR, q < 5%*) was used to consider the multiple comparisons correction. Furthermore, post-hoc analysis with Tukey’s Honest Significant Difference (HSD) was implemented to clarify the multiple comparison among various episodes.

### Cognition decoding for functional reorganization

We then further estimated the cognition relevance of functional reorganization patterns among different episodes of bipolar disorder using NeuroSynth (Yarkoni et al., 2011). We used twenty cognition maps from previous study (Margulies et al., 2016) and correlated them with functional gradient difference maps between BD episodes and HC via Pearson’s correlation. Finally, we visualized cognition terms according to these correlation values.

### Information exchange in functional reorganization regions across various episodes of bipolar disorder

To further examine the directional functional flow on reorganization regions, we employed regression dynamic causal modeling (rDCM) to parcellation preprocessed time sequence of each subject. rDCM is a computationally efficient framework expanding upon from classical dynamic causal modeling (DCM) framework. Compared with the original CDM, rDCM was designed to be more data-driven and implemented linear hemodynamic response function and mean-field approximation among various regions.A more detailed description of rDCM could be found elsewhere (Frässle et al., 2018, Frässle et al., 2017). Specifically, we fit parcellation-level time series from preprocess rs-fMRI via TAPAS ( https://www.tnu.ethz.ch/de/software/tapas) to obtain whole-brain effective connectome estimation (200×200 matrix) for each individual. To simplify directional information of resting state on the whole cortex, we estimated the weighted outward and weighted inward degree, reflecting outward and inward signal flows from specific regions respectively. To simplify the interpretation of cortical flow across functional reorganizatio regions, we further averaged the weighted outward and inward degree in previous significant regions in sensory regions and association regions respectively and compared them via *two-sample ttest*.

### functional integration and separation of cortex-wide connectome

We applied the Nested Spectral Partitioning (NSP) based on the eigenmodes method to detect hierarchical modules as well as quantify capacity for functional segregation and integration in FC networks. Differing from clustering and modularity-maximization methods, NSP method is based on a physical principle where regions with the same eigenvector sign are considered cooperatively activated, and regions with different signs are considered oppositely activated (R. Wang et al., 2022). Specifically, using spectral graph theory, the Laplacian matrix of whole-brain functional connectivity matrix is computed, and its eigenvalues and eigenvectors are utilized to perform an initial partition of the whole-brain functional network. This spectral partitioning minimizes the cut edges between regions, defining the initial functional modules. Based on this initial partitioning, each subnetwork is further partitioned in a nested manner, recursively continuing this process until a predetermined partitioning scale or other termination criteria are met. At each level of partitioning, the strength of connections within each subnetwork (reflecting segregation) and between subnetworks (reflecting integration) are calculated (R. Wang et al., 2022). These metrics quantify the segregation and integration of the brain’s functional network at different hierarchical levels. It is noteworthy that the NSP method is influenced by the length of the signal used to construct functional connectivity. However, in this study, the resting-state data collection duration for each subject was consistent.

Finally, we evaluated the segregation and integration coefficients of whole-brain functional connectivity for each individual and used a one-way-anova and post-hoc analysis to compare those coefficients’ differences between the disease group and the healthy control group with age and sex as covariates.

### Functional reorganization and clinical symptom association

To explore clinical symptoms of functional reorganization, we further linked functional gradient values to clinical symptom estimation. We averaged functional gradient values in previous significant regions in sensory regions and association regions respectively. Then, we calculated Pearson’s correlation between gradient alteration in BipM,rBD, BipD with YMRS,HRSD,HAMA, and PDQ respectively.

### Molecular mechanism of functional reorganization Functional reorganization and Receptor maps

Receptor densities were assessed through the utilization of PET tracer investigations encompassing a total of 18 receptors and transporters spanning nine neurotransmitter systems. This data, recently shared by Hansen and colleagues (https://github.com/netneurolab/hansen_receptors) (Hansen et al., 2022a). The neurotransmitter systems include dopamine (D1, D2 DAT), norepinephrine (NET), serotonin (5-HT1A, 5-HT1B, 5-HT2, 5-HT4, 5-HT6, 5-HTT), acetylcholine α4β_2_, M1, VAChT), glutamate (mGluR5), GABA (GABAa), histamine (H3), cannabinoid (CB1), and opioid (MOR)(Markello et al., 2022). Volumetric PET images were aligned with the MNI-ICBM 152 nonlinear 2009 (version c, asymmetric) template. These images, averaged across participants within each study, were subsequently parcellated into the Schaefer 200 template. Receptors/transporters exhibiting more than one mean image of the same tracer (5-HT1B, D2, VAChT) were amalgamated using a weighted average (Hansen et al., 2022a).

### Functional reorganization and Cellular maps

Here, we further correlated patterns observed in three episodes of bipolar disorder with 24 cellular maps from a previous study (X.-H. Zhang et al., 2023). The molecular signature profiles of all cell classes were constructed from snDrop-seq samples provided by (Jorstad et al., 2023). Then cell type fractions were deconvolved from microarray samples downloaded from Allen Human Brain Atlas (AHBA; http://human.brain-map.org/) (Shen et al., 2012). The 24 cell types are Lamp5, Pax6, Vip, Sncg, Lamp5, Lhx6, L5ET, L5/L6 NP, L6 CT, L6b, Astro, VLMC, Endo, Micro/PVM, Oligo, OPC, L2/3 lT, L6 lT Car3, L4 lT, L6 lT, L5 lT, Chandelier, Pvalb, Sst, and Sst Chodl, as detailed in (X.-H. Zhang et al., 2023).

### Null model

In our study, we aimed to explore the topographic correlations between functional reorganization patterns and other notable features. To infer these correlations, we implemented a null model that systematically disrupted the relationship between two topographic maps while maintaining their spatial autocorrelation (Markello & Misic, 2021). Initially, we shuffled receptor maps or cellular maps and examined their relationship with functional reorganization patterns . The resulting spatial coordinates provided the basis for generating null models through a process of randomly sampled rotations and reassignment of node values based on the nearest resulting parcel (Hansen et al., 2022b). This process was iterated 1000 times. Importantly, the rotation was first applied to one hemisphere and then mirrored onto the other. The threshold value was determined by the 95th percentile of shuffling occurrence frequencies derived from spatial null models.

## DATA AVAILABILITY

The clinical data could be accessed according to reasonable requests for corresponding authors. The raw fMRI data and MRI data for HCP was available on https://db.humanconnectome.org/. Heritability analyses were performed using Solar Eclipse 8.5.1b (https://www.solar-eclipse-genetics.org), Neuromap was available on (https://netneurolab.github.io/neuromaps/usage.html).

## CODE AVAILABILITY

Code will be available on https://github.com/Laoma29/Publication_codes.

## ACKNOWLEDGMENTS

Xiaobo Liu, Siyu Long, Jiadong Yan and Ke Xie are supported by the China Scholarship Council. Bin Wan is supported by International Max Planck Research School on Neuroscience of Communication: Function, Structure, and Plasticity (IMPRS NeuroCom), Graduate Academy Leipzig, and Mitacs Globalink Research Award. ZQL acknowledges support from the Fonds de Recherche du Qu\’ebec -- Nature et Technologies (FRQNT). Yujun Gao is supported by the Health of Hubei Province Scientific Research Project under Grant 2020Cfb512, and by the Mental Health Research Institute of Three Gorges University: YCXL-23-11.

## COMPETING INTERESTS

No competing interests among the authors.

## Supplementary Materials

### Research Participants Collection Procedures and Data Quality Control

The study initially recruited a larger cohort, but ultimately excluded 92 participants due to specific criteria ensuring the integrity of the dataset. In the BipD group, 9 participants were excluded after being diagnosed with major depressive disorder during follow-up, 1 due to the presence of organic brain disease, 2 for inability to complete the scan, 4 for excessive head motion, and 4 due to the presence of metal braces. The BipM group saw the exclusion of 8 participants for excessive head motion, 2 for misdiagnosis during follow-up, 4 with comorbid gender identity disorder, 2 with organic brain disease, and 8 who were unable to complete the scan. In the rBD group, 2 participants were excluded following a follow-up diagnosis of major depressive disorder.In the rBD group, saw the exclusion of 6 participants for excessive head motion. Within the HCs group, exclusions comprised 4 participants with metal braces, 4 with head motion exceeding 2mm who did not consent to rescanning, and 15 with a significant family history of psychiatric or neurological disorders among first-degree relatives. After these exclusions, the study included 42 BipD patients, 38 BipM patients, 37 rBD patients, 38 mBD patients,and 35 HCs, with all BipM patients completing follow-up through various methods (see Figure. S1 for details). This meticulous selection process underscores the importance of stringent inclusion criteria and rigorous data quality control in ensuring the reliability and validity of the research findings.

**S-Figure.1.**
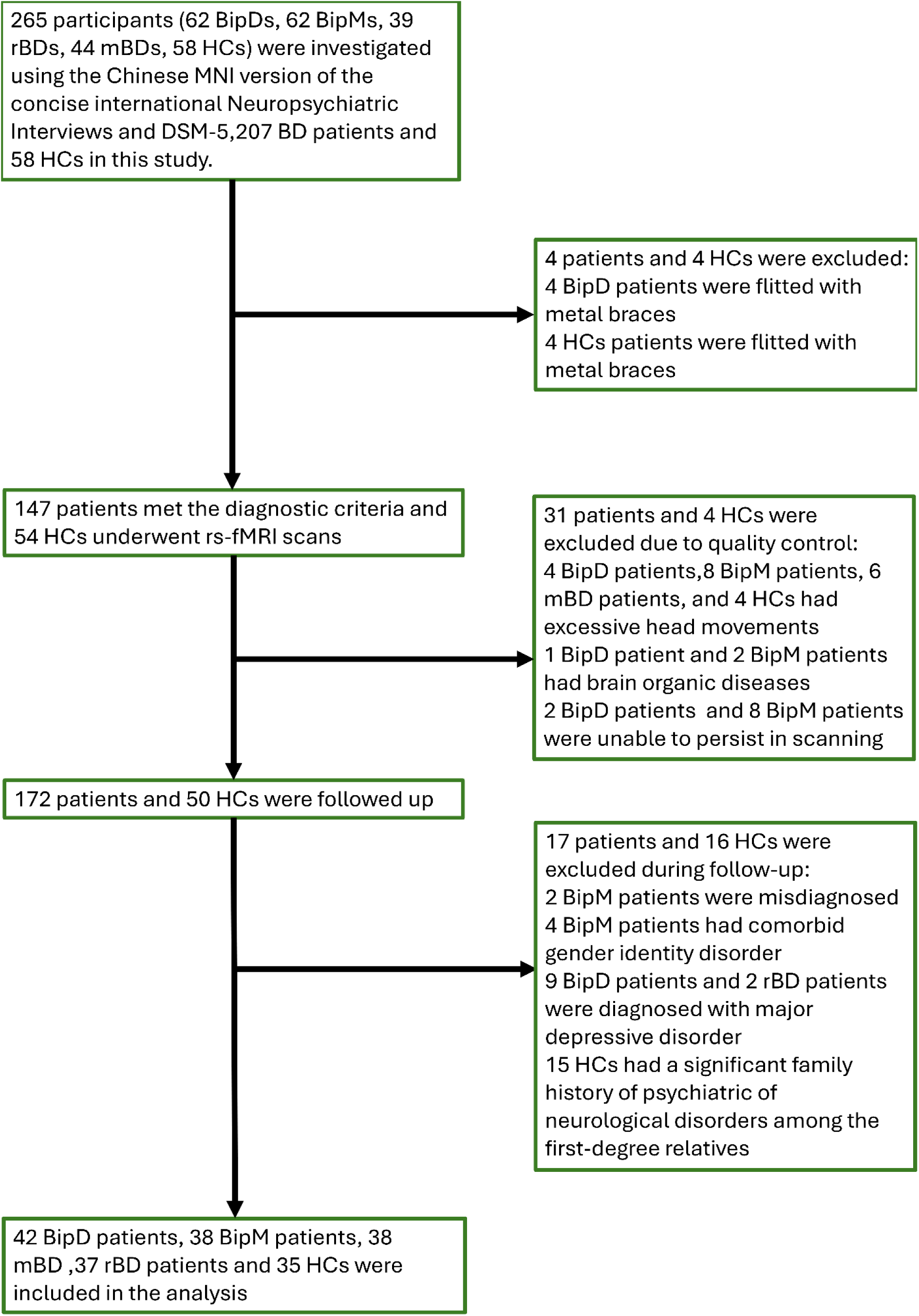
The research participants collection procedures and data quality control.

**S-Figure.2.**
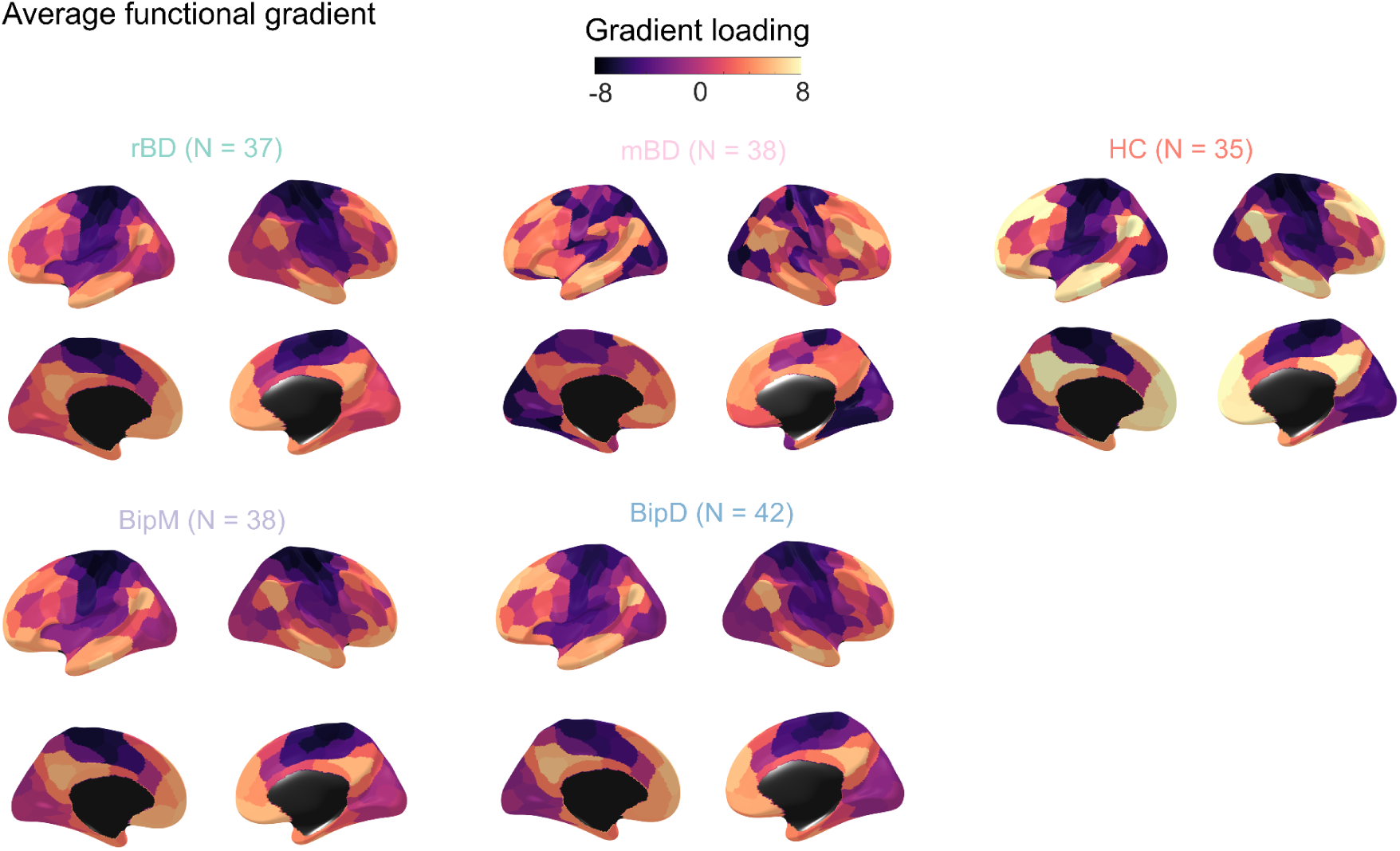
Average functional gradient map for rBD, mBD, BipM, BpD and HC.

**S-Figure.3.**
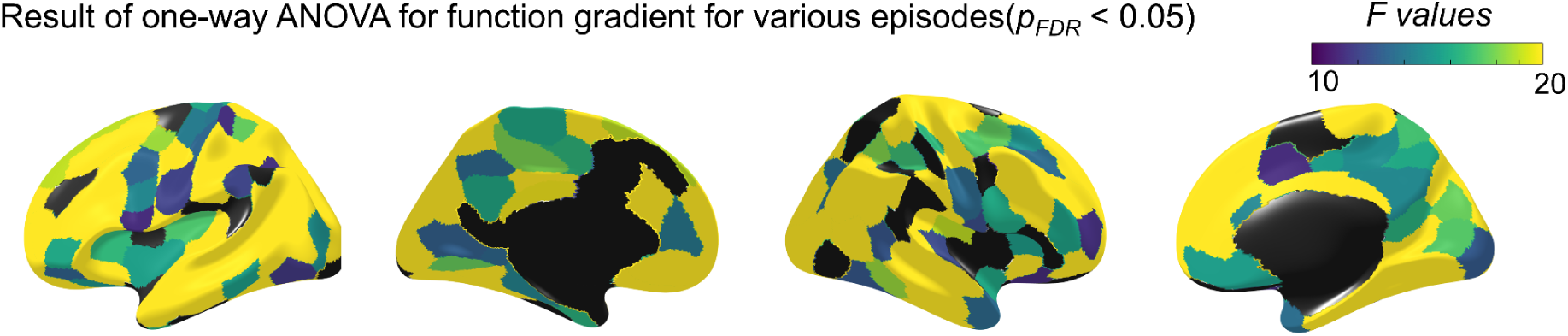
F value map across various episodes (*p_FDR_< 0.05*).

**S-Figure.4.**
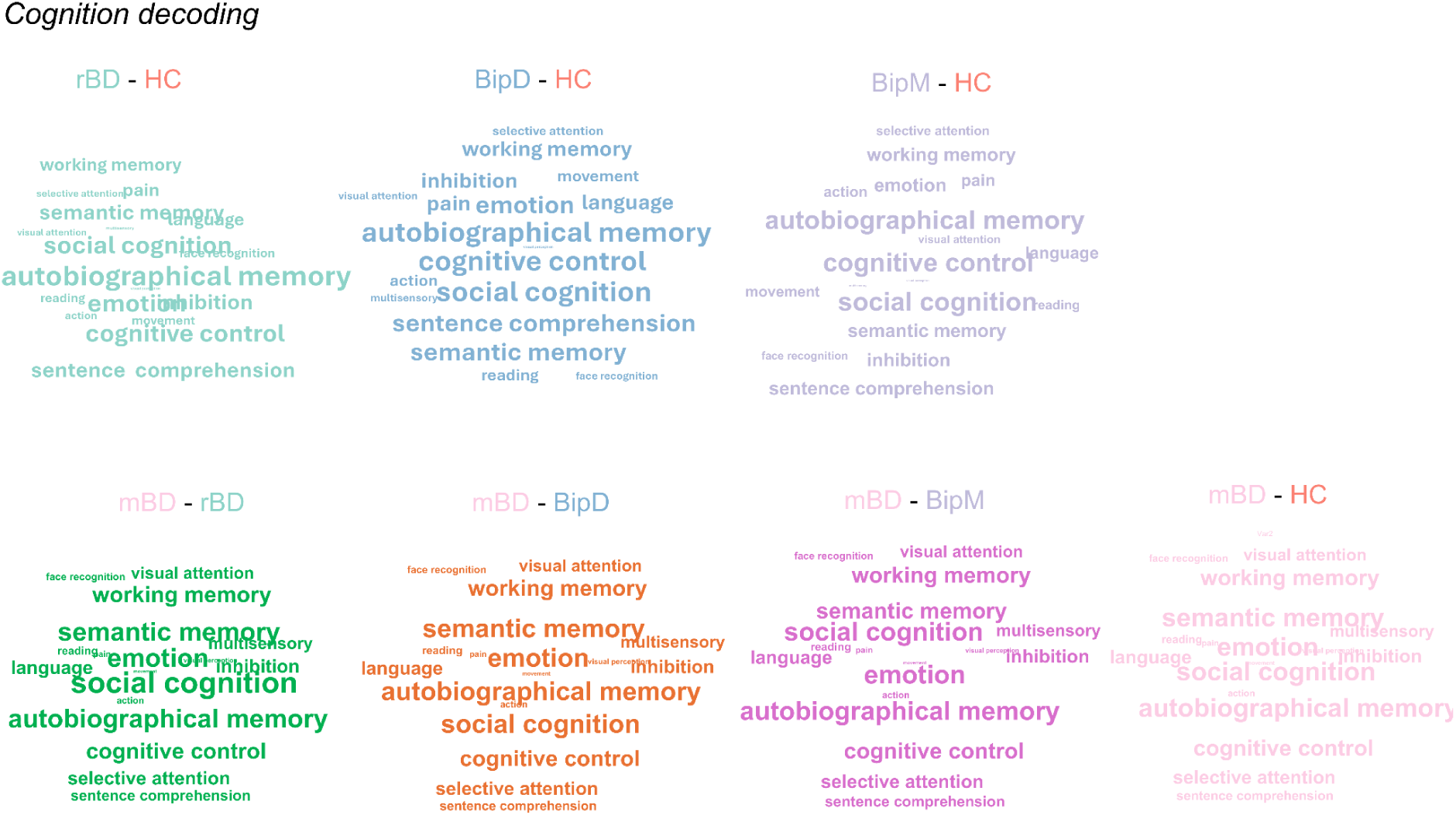
Cognition decoding of functional reorganization across various episodes.

**S-Figure.5.**
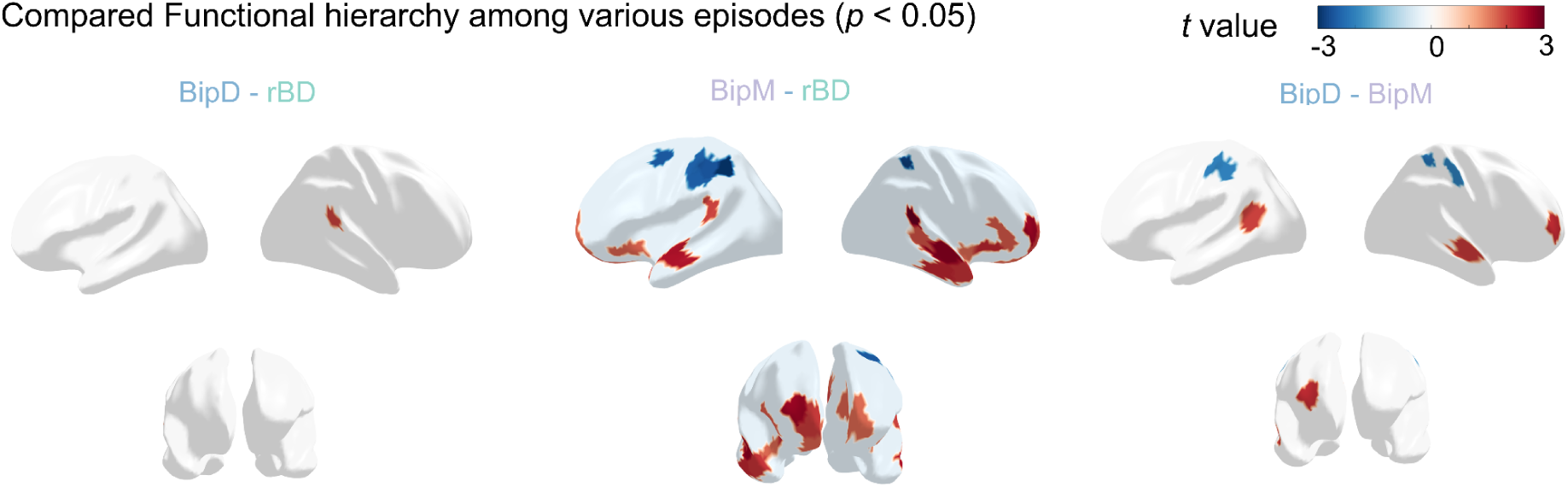
The comparison of functional gradients across rBD, BipM and BipD (*p< 0.05*).

**S-Figure.6.**
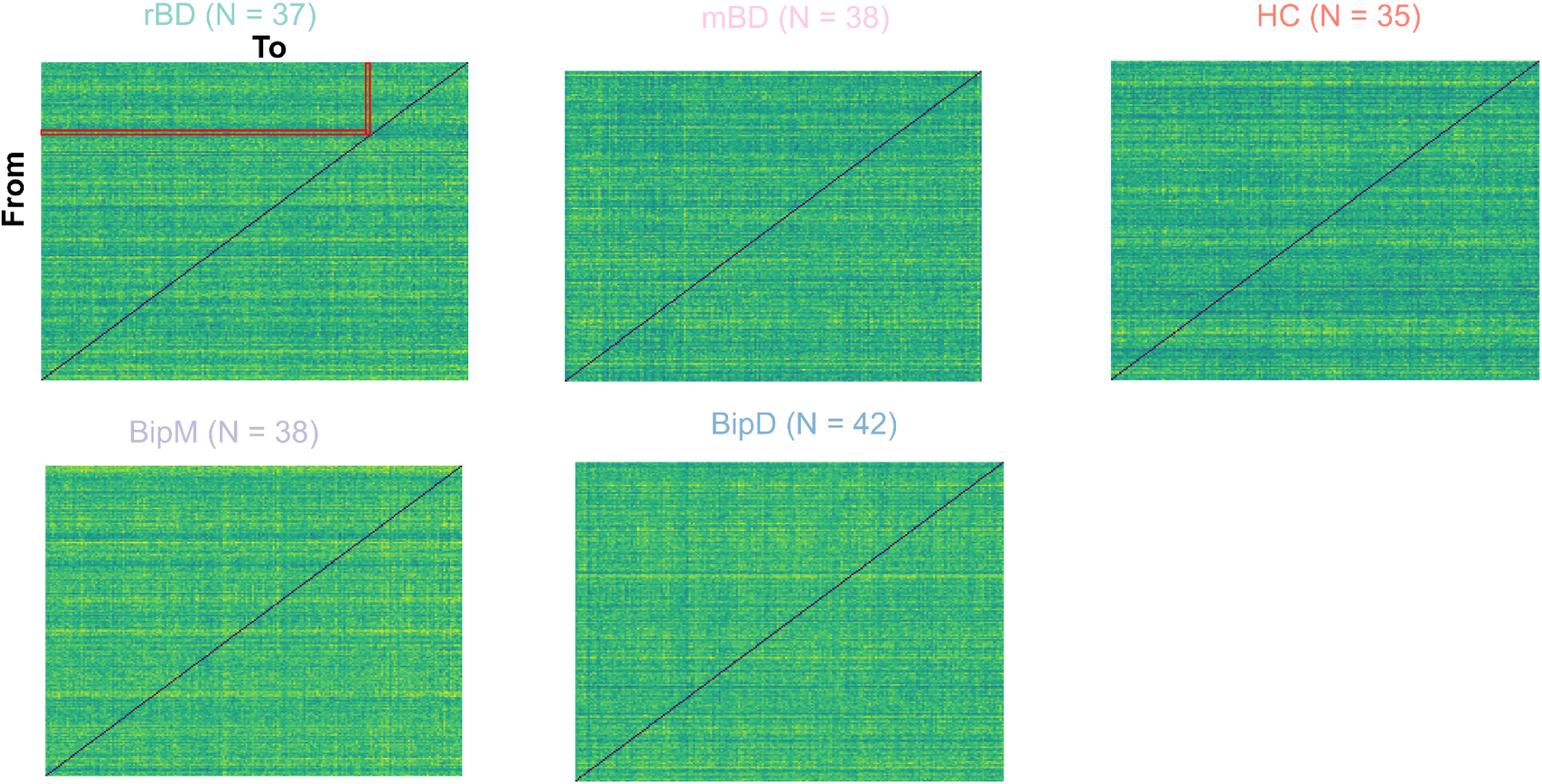
The averaged information exchange matrices based on rDCM across rBD, mBD, BipM, BipD and HC.

**S-Figure .7.**
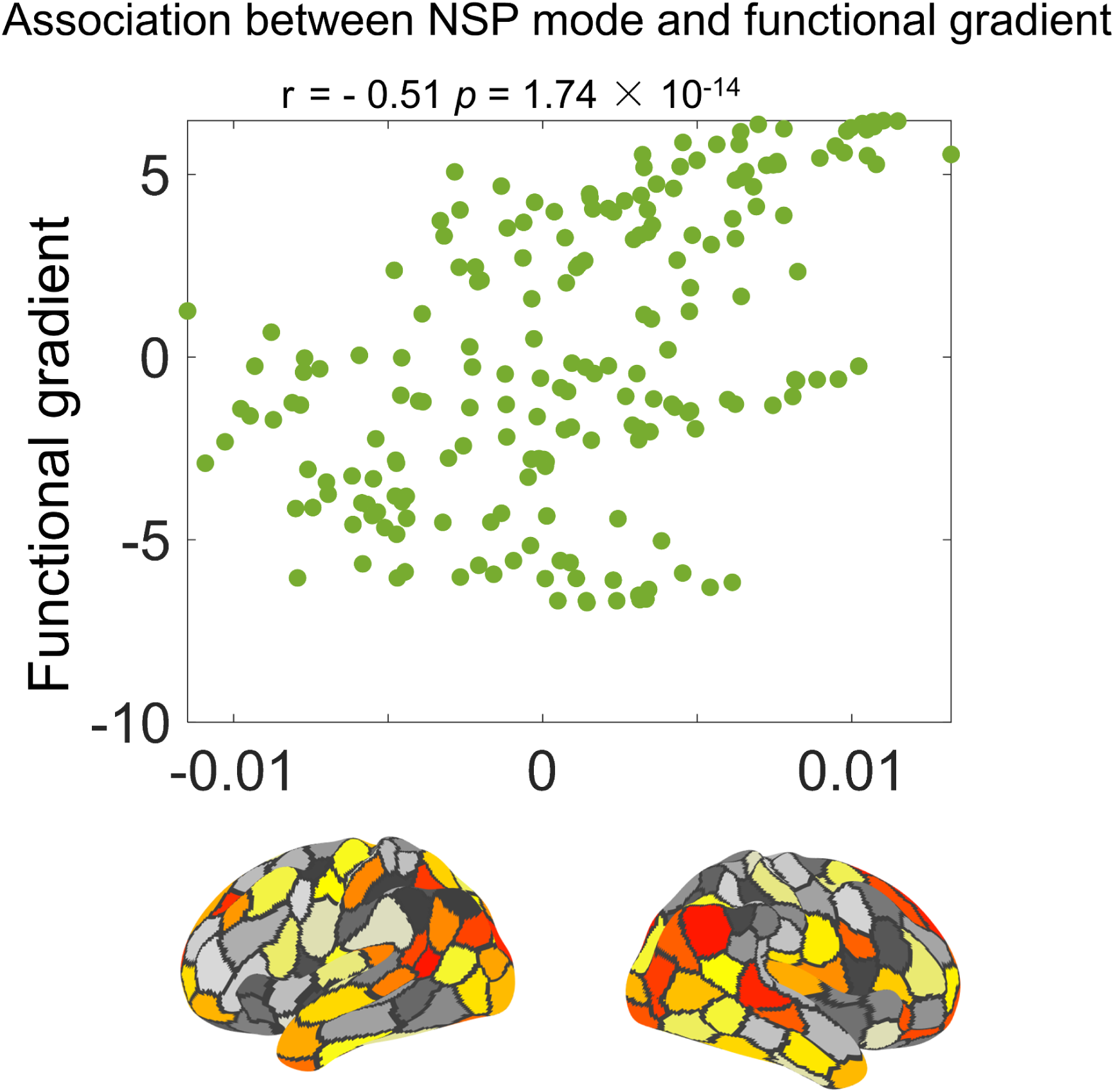
The correlation between first NSP mode and functional gradient pattern (*r = 0.51, p = 1.74× 10^−14^*)

**S-Figure .8.**
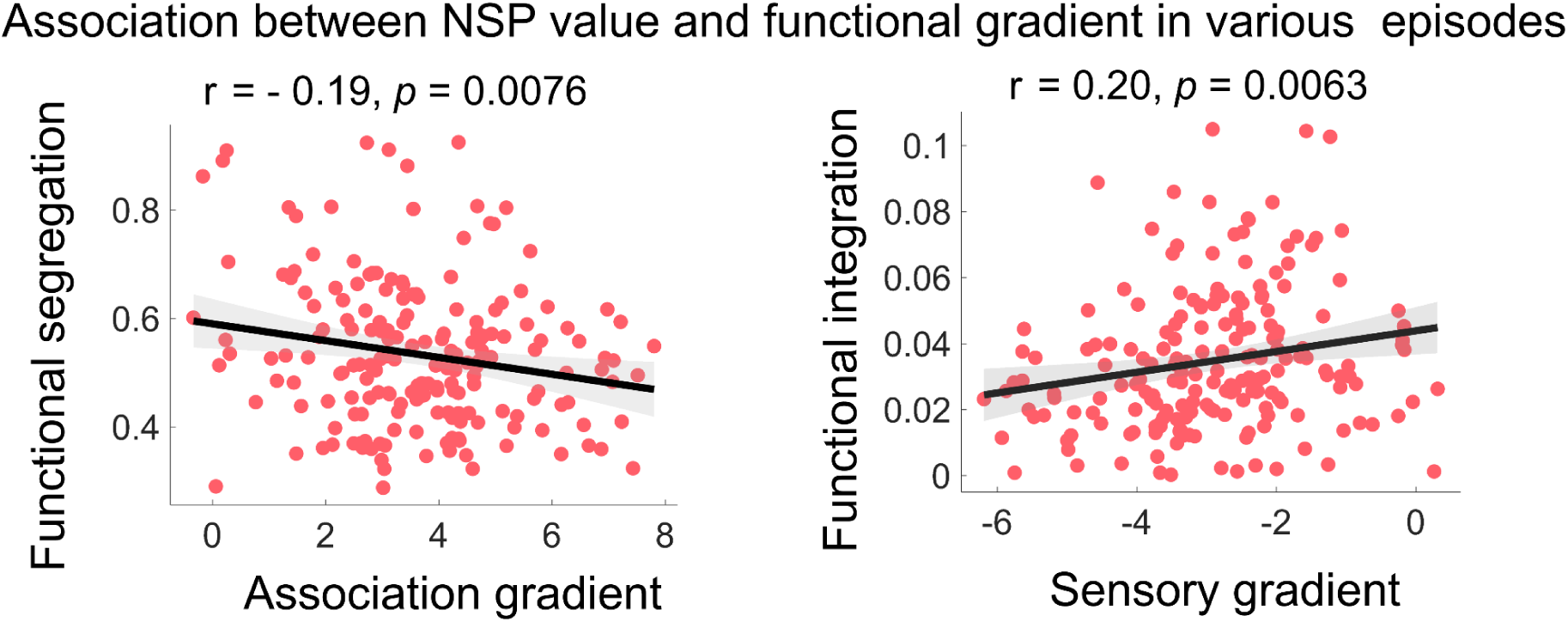
Functional integration was positively associated with sensory reorganization (*r = - 0.19, p = 0.0076*), Functional separation with association reorganization regions *(r = 0.20, p = 0.0063*).

**S-Figure .9.**
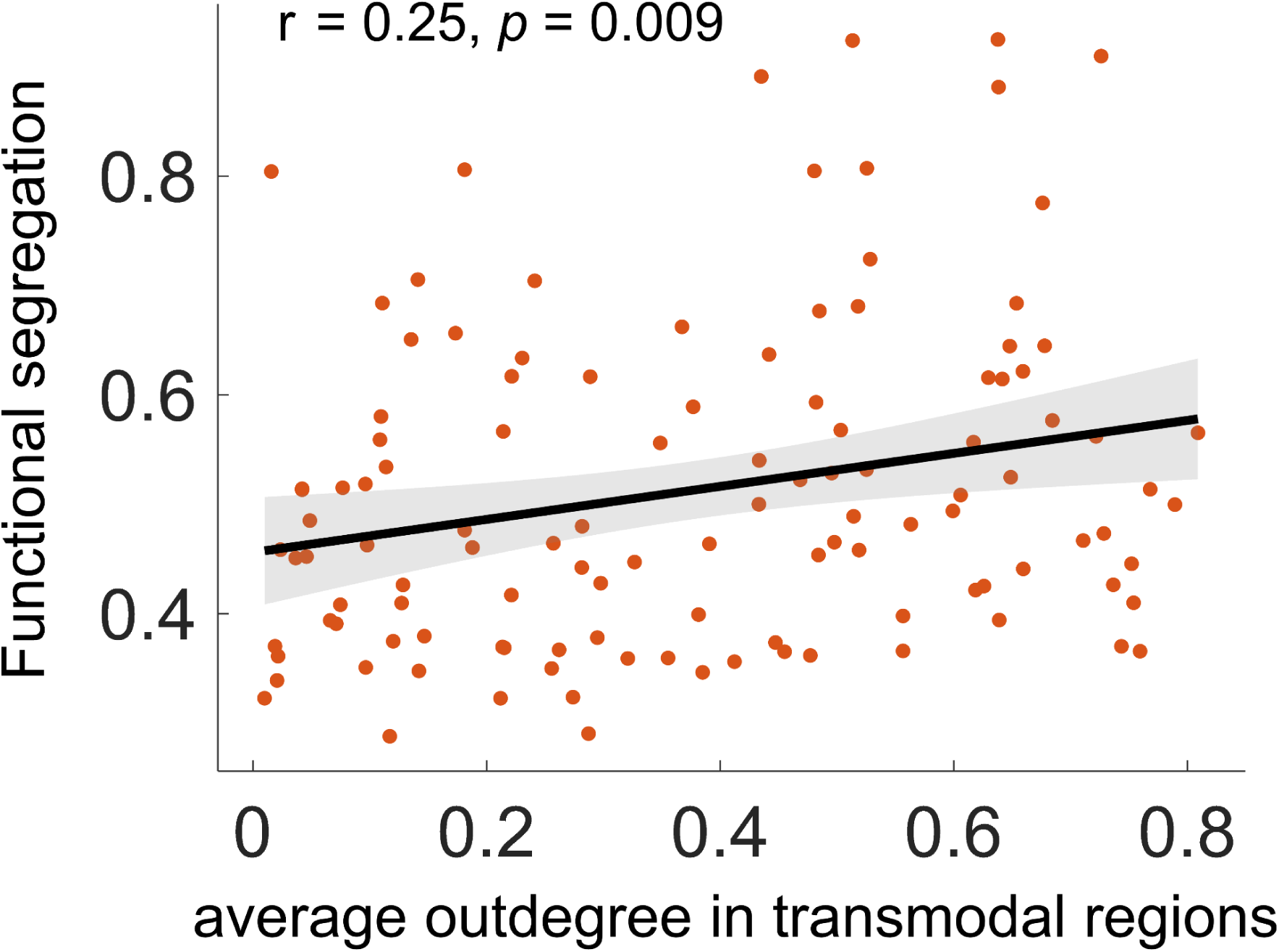
The correlation between functional segregation and average outdegree in association regions ( *r = 0.25*, *p = 0.009*)

**S-Figure .10.**
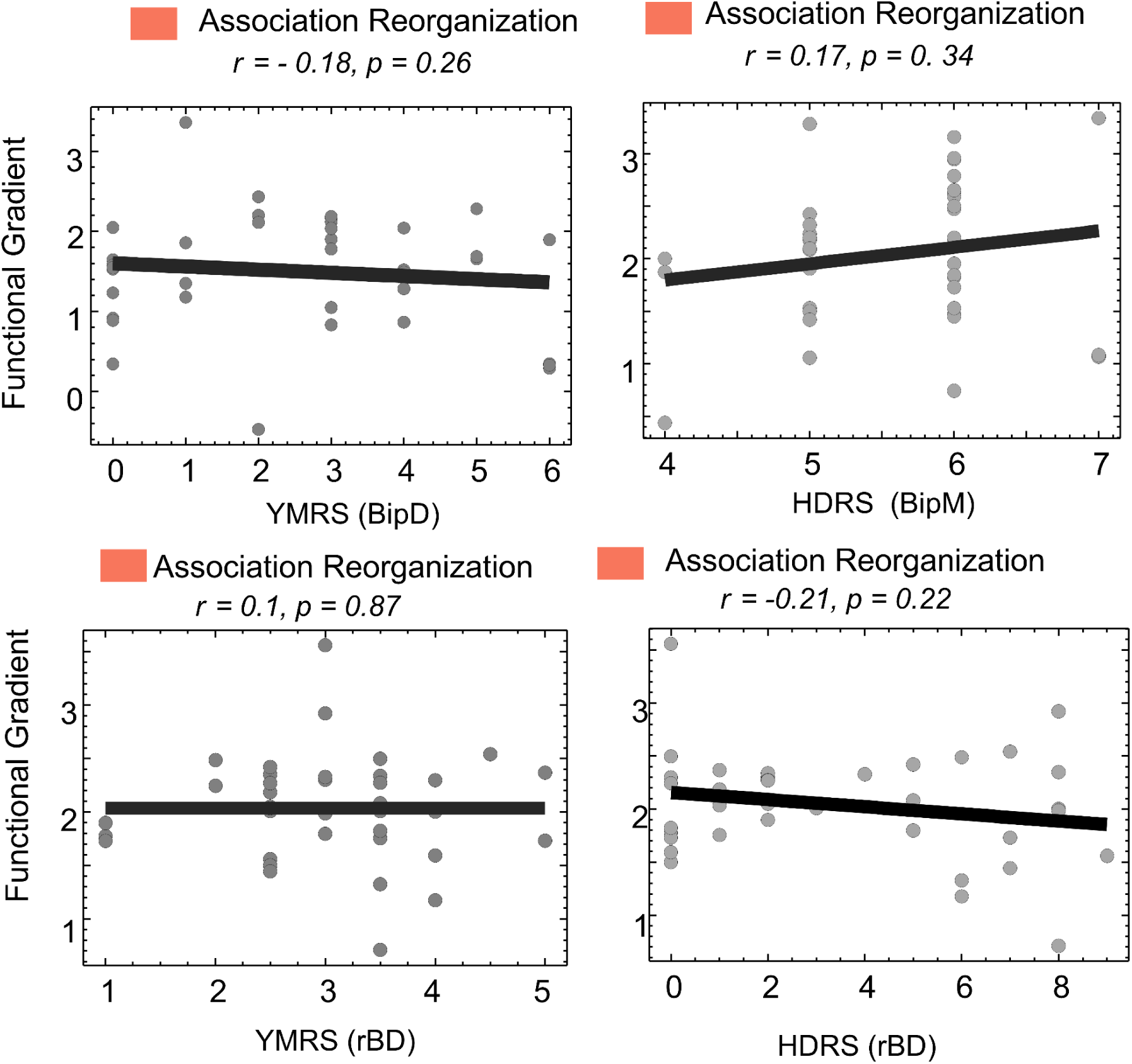
The correlation between clinical symptom and functional reorganization in association regions in BipD( *r = - 0.18*, *p = 0.26*), BipM( *r = 0.17*, *p = 0.34*) and BipD ( *r = 0.1*, *p = 0.87* and *r = - 0.21*, *p = 0.22*).

## Notes

### Competing Interest Statement

The authors have declared no competing interest.

### Funding Statement

This study did not receive any funding

### Author Declarations

This study was conducted following ethical approval from the Ethics Committee of Renmin Hospital of Wuhan University (Ethics Approval Number: WDRY22022-K195).

### Summary of Updates

We update new version of dataset.

